# Synaptic inhibitory dynamics drive benzodiazepine response in paediatric status epilepticus

**DOI:** 10.1101/2023.08.23.23294456

**Authors:** Tommaso Fedele, Richard J. Burman, Anne Steinberg, Giorgio Selmin, Georgia Ramantani, Richard E. Rosch

## Abstract

Paediatric status epilepticus (SE) is a medical emergency associated with significant morbidity. Benzodiazepines (BZPs) are the current first-line treatment, but do not work in over a third of children presenting with SE. Animal studies have shown that SE can cause changes in synaptic inhibition signalling which can ultimately lead to BZPs becoming ineffective. However, the relevance of these mechanisms in paediatric patients with SE remains unknown. To test this hypothesis, we combine clinical EEG recordings with dynamic causal modelling (DCM). This approach allows model-based inference of cortical synaptic coupling parameters based on EEG recorded across distinct oscillatory states. Our DCM revealed that dynamic changes in inhibitory synaptic coupling explain differences in EEG power spectra associated with BZP-treatment responsiveness, and guide the transition from ictal to interictal state. Furthermore, *in silico* simulations demonstrate that there are alternative routes to seizure termination even in cortical circuit models unresponsive to the BZPs. Together, our findings confirm that alterations in synaptic inhibition underlie BZP response during paediatric SE. More broadly this work further demonstrates the utility of computational modelling to validate insights from basic science in clinically accessible recordings in neurological disorders characterised by abnormal brain states.

## Introduction

Paediatric status epilepticus (SE) remains a pertinent clinical challenge. This state of prolonged and unrelenting seizure activity is a medical emergency as it is associated with significant morbidity in children^1^ as well as being a burden on healthcare resources.^2,3^ Benzodiazepines (BZPs), available in different subtypes and formulations, are the preferred first-line treatment due to their ease of administration, fast onset time and, in most cases, effectiveness in terminating SE.^4–6^ However, in up to 32% of children presenting with SE, BZPs do not work.^7^ The lack of response to BZP in these children adds complexity to their management, often requiring additional medications and an increased demand on PICU services which is often limited especially in lower-middle-income countries.^8,9^

Animal studies have demonstrated that underlying the resistance to BZPs are SE-induced changes in synaptic physiology that alter the main target of these drugs, the GABA-A receptor (GABA_A_R). These chloride (Cl^-^)-permeable channels play a principal role in mediating inhibitory signalling in the brain and helping restrain aberrant electrical activity such as seizures.^10,11^. During SE-like activity, there is a cascade of changes that affect the GABA_A_R which culminate in a loss of intact synaptic inhibition.^7,12^

While these SE-dependent changes in synaptic inhibition appear robust in animal models, their relevance in the pathophysiology of BZP resistance in paediatric patients with SE is unknown. It is here that computational modelling may provide a useful way of translating between the experimental findings and neurophysiological recordings routinely gathered from patients. Modelling the activity of recurrently coupled excitatory and inhibitory neuronal populations is already established for understanding different types of seizure activity, including SE.^13,14^ One such computational framework is dynamic causal modelling (DCM) which has demonstrated utility in studying the hierarchical structure of state-transitions in neuronal population activity during epileptic seizures.^15–18^ An appealing feature of DCM is that it can infer regional disease-related changes in the cortex, specifically excitatory and inhibitory synaptic transmission between different cortical layers believed to be generating the EEG signal.^19,20^ In this way, DCM offers an opportunity to study how changes in synaptic dynamics contribute to disease pathophysiology, which is highly relevant in the context of BZP-resistant SE.

Here, we combine scalp EEG recordings with DCM to explore the mechanisms underlying BZP resistance in paediatric SE. This is a unique cohort for whom drug administration during SE was captured by continuous EEG monitoring as part of routine clinical care. Through our analysis of this dataset, we demonstrate how changes in synaptic inhibition differentiate children who are unresponsive to BZPs. We also reveal that patients who are resistant to BZPs appear to have both pre-existing and acquired electrographic features that appear to affect their sensitivity to BZPs.

## Materials and methods

### Patient selection

This study used retrospective anonymised patient data and was approved by the local ethics committee (Kantonale Ethikkommission Zürich, KEK-ZH PB-2020-02580). A consent waiver was granted. We retrospectively identified paediatric patients with SE (<18 years of age) who underwent scalp EEG recordings between July 2008 and February 2020 at the University Children’s Hospital Zurich. Our inclusion criteria were electroclinical diagnosis of status epilepticus, and administration of an antiseizure medication (ASM) during the EEG recording. Neonatal patients (<1 month of age) were excluded.

### EEG acquisition and analysis

Clinical EEG recordings (21 electrodes, international 10–20 electrode layout, 256-Hz sampling frequency, 1-70 Hz band pass) were visually reviewed and annotated by two EEG reviewers with relevant training in epileptology or clinical neurophysiology (G.S. and G.R.) and discrepancies were resolved by consensus. Annotations included characterisation of electroclinical features and characterisation of ASM treatments (Supplementary Table 1). To evaluate the effect of ASM on the EEG, we identified five-minute artifact-free EEG intervals preceding and following ASM administration: pre-medication intervals, terminating one minute before drug administration, and post-medication intervals, starting six minutes after drug administration in non-responders and one minute after the termination of the continuous status epilepticus or the last seizure in intermittent status epilepticus in responders. Six minutes was an empirical choice guided by an average response time to BZPs of 340±40 s (mean ± Standard Error) in our cohort.

### Statistical analysis

To quantify group-level differences in the pre- and post-medication EEG spectra of responders and non-responders to BZP we used a repeated-measures (rm) ANOVA using the factors treatment time (pre- vs. post-medication) and treatment efficacy (responders vs. non-responders). Post-hoc analysis was performed with paired (within subjects) and unpaired (between groups) Wilcoxon’s test (Bonferroni correction, *n* = 6 (4 groups), significance at uncorrected *P* = 0.0083).

### Dynamic causal modelling (DCM)

In order to infer synaptic parameters governing neuronal populations’ ongoing oscillatory activity, we used a DCM for cross-spectral densities in electrophysiological data (SPM12). Here, we fitted single-source DCMs consisting of four coupled neural masses (corresponding to the canonical microcircuit) to dimensionality-reduced single-channel representations of the EEG activity pre- and post-medication for each patient individually. To identify which synaptic parameters are consistently changed by the effect of BZP administration, we used a parametric empirical Bayesian approach^21^, which allows inferring effects across individual DCMs. This approach is described in more detail in the Supplementary Methods.

### Simulations of effects of individual synaptic parameters

The DCM framework allows to extract optimal values of synaptic parameters fitting a given power spectrum. Conversely, we can simulate the effect of altering parameter values on the power spectrum. We devised simulations aimed to reproduce in silico the group level effects extrapolated by the second level DCM. Additionally, we performed a fully comprehensive parameter screen across the 14 synaptic parameters (Supplementary Fig. 1). In this way we identified parameters that allow for non-responders to ‘escape’ the ictal power spectral patterns.

### Data availability

The code used to produce the results presented in this manuscript is available at https://github.com/tommytommy81/SE_DCM_code.

## Results

### Differences in spectral features characterise variable responses to BZPs in paediatric SE

We analysed the EEG recordings of paediatric patients with SE treated with BZPs and other ASMs (Fig. 1A-B, Supplementary Table 1). Among patients treated with BZP, those who showed a clear resolution of the SE on visual EEG interpretation were labelled responders, while those in which SE persisted were labelled non-responders (Fig. 1C). The modulation of the EEG was reflected in the power spectra of extracted ‘virtual’ local field potentials (vLFP) used for DCM analysis which revealed a significant effect of BZP treatment across responders vs non-responders to BZP (Fig. 4D). Post-hoc analysis revealed significant differences between responders and non-responders in pre-medication delta power. In responders the pre- to post-medication delta-alpha range spectral power differed significantly, whereas in non-responders no difference was detected.

**Figure 1:**
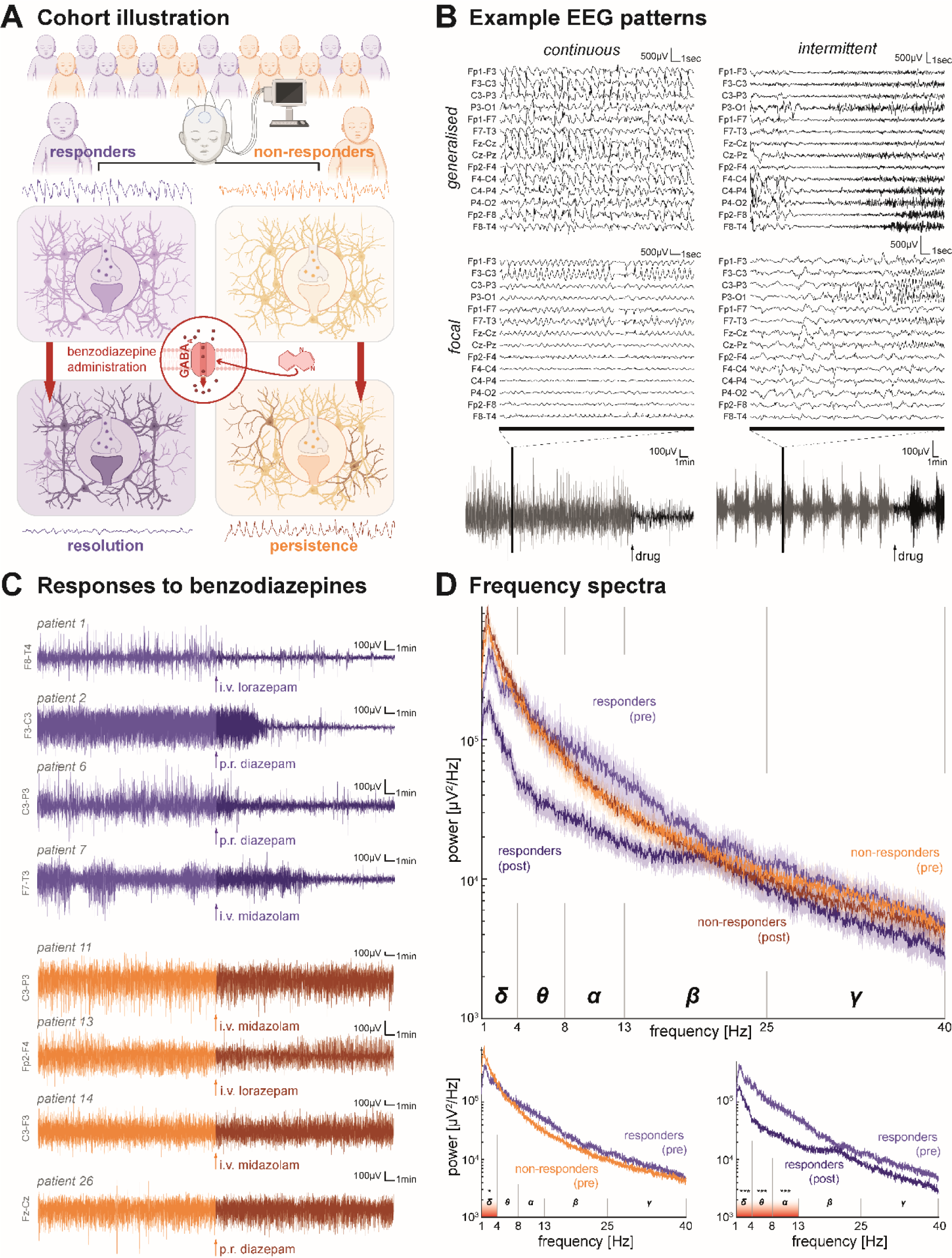
Differences in spectral features characterise variable responses to benzodiazepines (BZPs) in paediatric status epilepticus. (**A**) Schema of the study design in which EEG recordings were acquired from a cohort of patients (*n* = 26) who received BZPs during an episode of status epilepticus (SE, *n* = 17). BZPs are GABA-A receptor (GABA_A_R) allostatic modulators and expected to enhance synaptic inhibition. Patient whose SE was terminated by BZPs were labelled ‘responders’ (purple, *n* = 8) whilst those whose SE persisted after BZP treatment were labelled ‘non-responders’ (orange, *n* = 9). (**B**) Raw example EEG traces of different electrographic presentations of paediatric SE with different spatial (i.e., generalised vs focal) and temporal (i.e., continuous vs intermittent) features that were included in the analysis. Bottom panel illustrated zoomed-out view of single electrode recording and demonstrates period before (grey) and after (black) BZP administration (arrow). (**C**) Raw example EEG traces demonstrating both responsive patients (top panel, purple) and non-responsive patients (bottom panel, orange) before (lighter shade) and after (darker shade) different administrations and formulations of BZPs. (**D**) Spectral analysis of EEG traces reveals difference in power spectra between responders and non-responders before (‘pre’) and after (‘post’) BZP administration in the theta band (dark line illustrates mean, shade illustrates the SEM, F= 1.85, *P* =0.02, two-way repeated measures *ANOVA*). Across all patients, before BZP administration (bottom left panel) there was a significant difference in delta band power between responders and non-responders (responders: 2.74e5 ± 3.1e4 µV^2^/Hz vs non-responders: 4.27e5 ± 4.5e4 µV^2^/Hz, *P_corr_* =0.031, *Wilcoxon’s test*). In responders, BZP administration significantly changed the spectra in the delta (responders-pre: 2.74e5 ± 3.1e4 µV^2^/Hz vs responders-post: 1.12e5 ± 1.27e4 µV^2^/Hz, *p_corr_* <0.001, *Wilcoxon’s test*), theta (responders -pre: 1.37e5 ± 1.53e4 µV^2^/Hz vs responders-post: 3.72e4 ± 4.16e3 µV^2^/Hz, *p_corr_* <0.001, *Wilcoxon’s test*), alpha (responders-pre: 7.06e4 ± 7.9e3 µV^2^/Hz vs responders-post: 2.3e4 ± 2.57e3 µV^2^/Hz, *P_corr_*<0.001, *Wilcoxon’s test*) frequency bands. ‘i.v.’, intravenous; ‘p.r.’, per rectal; ‘δ’, delta band (1-4 Hz); ‘θ’, theta band (4-8 Hz); ‘α’, alpha band (8-13 Hz); ‘β’, beta band (14-30 Hz); ‘γ’, gamma band (>30 Hz); ‘***’, *P* < 0.001; ‘BZP’, benzodiazepines; GABA-A, γ-aminobutyric acid receptor type A.

### Dynamic causal modelling enables estimation of changes in synaptic parameters

Next, we fitted the vLFP power spectra with a neural mass model through DCM. From the clinical EEG (Fig. 2A) we extracted the vLFP best representing SE (Fig. 2B). The core component of the DCM is the canonical micro-circuit^22^ (Fig. 2C). DCM can be fitted to the vLFP power spectra for different time windows, and allow for synaptic parameters to be extracted (Fig. 2D). These parameters are functionally grouped here as time constants as well as excitatory and inhibitory coupling (Fig. 2E). For subsequent group-level analysis, we included a single pre- and post-medication time window for each patient. This yielded two ‘first level’ DCMs for each patient, fitting the pre- and post-medication power spectra (Fig. 2F) through model inversion, providing estimates of synaptic parameters (Fig. 2G).

**Figure 2:**
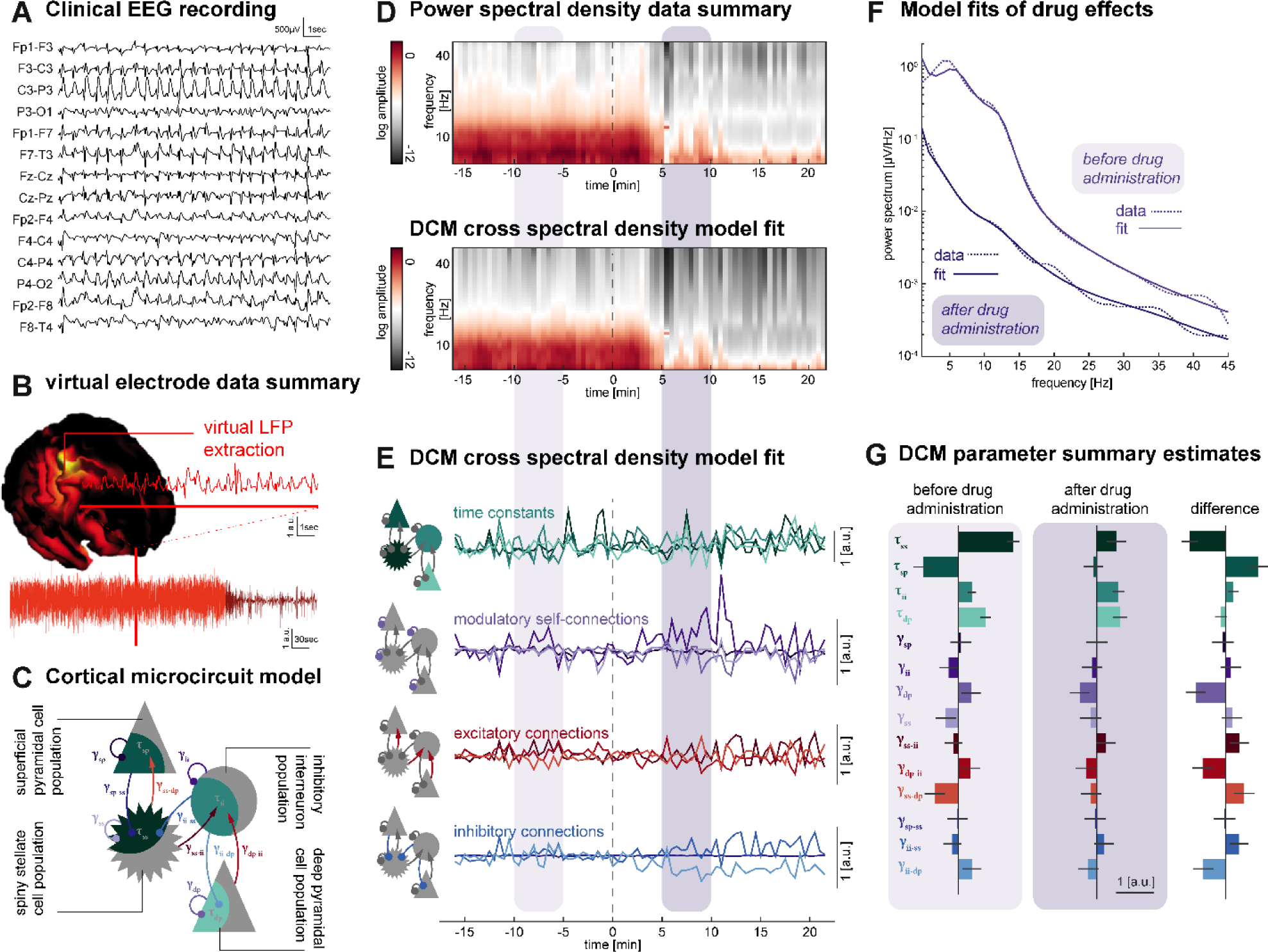
Dynamic causal modelling enables estimation of changes in synaptic parameters from clinical recordings of paediatric status epilepticus. (**A**) Example raw EEG trace from a single patient (responder) which is then used to extract a virtual local field potential (LFP, **B**) that can then be used to study the effect of BZP administration (arrow). Red line highlights zoomed-in view of trace. (**C**) Schematic of canonical microcircuit model (CMC) of a cortical column, consisting of four coupled neural masses, representing layered populations capturing key aspects of intracortical connectivity and population dynamics. These include: superficial (supragranular) pyramidal neurons, spiny stellate excitatory interneurons, inhibitory GABAergic interneurons, and deep (infragranular) pyramidal neurons. Connections indicate synaptic connectivity parameters that can be excitatory (red), inhibitory (blue), or self-modulatory (purple). Neuronal activity is further characterised by time constants (green) governing the response of populations to input. These generative models are used to estimate the contribution of different excitatory and inhibitory synaptic parameters to observed EEG patterns (see Supplementary Table 2). (**D**) Time-resolved power spectral density data (30 s) from the virtual LFP before (top panel) and after being fitted by DCM (bottom panel). Five-minute epochs before (light purple) and after (dark purple) BZP administration (dashed line) were used in parameter analysis. (**E**) Time series of changes in cortical microcircuit physiological parameters including time constants (green), modulatory self-connections (purple), excitatory connections (red) and inhibitory connections (blue). (**F**) Spectral analysis of virtual LFP trace from five-minute epochs before (light purple) and after (dark purple) BZP administration highlighted in **D**. Empirically recorded (dashed lines) and DCM fitted data (solid lines) are superimposed for pre- and post-medication windows. (**G**) Summary of changes in cortical microcircuit physiological parameters before and after BZP administration for a single patient. BZP, benzodiazepine; CMC, canonical microcircuit; DCM, dynamical causal modeling; PSD, power spectral density; vLFP = virtual local field potential.

### Synaptic inhibition during SE differs between BZP responders and non-responders

We inferred which synaptic coupling parameters underly group-level differences in spectral power across pre- and post-medication time windows (within subjects), as well as across responders and non-responders (across subjects). For this we employed hierarchical DCMs, considering the effect of responsiveness, BZP treatment, and their interaction as a second-level design matrix (Supplementary Fig. 2). We formulated three main hypotheses regarding the between-subject effect of BZP responsiveness (Fig. 3A). For these, synaptic parameters in responders and non-responders either (1) differ during SE but are similarly altered by BZPs; (2) are similar during SE but are distinctly altered by BZPs; or (3) both differ in SE and are distinctly altered by BZPs.

**Figure 3:**
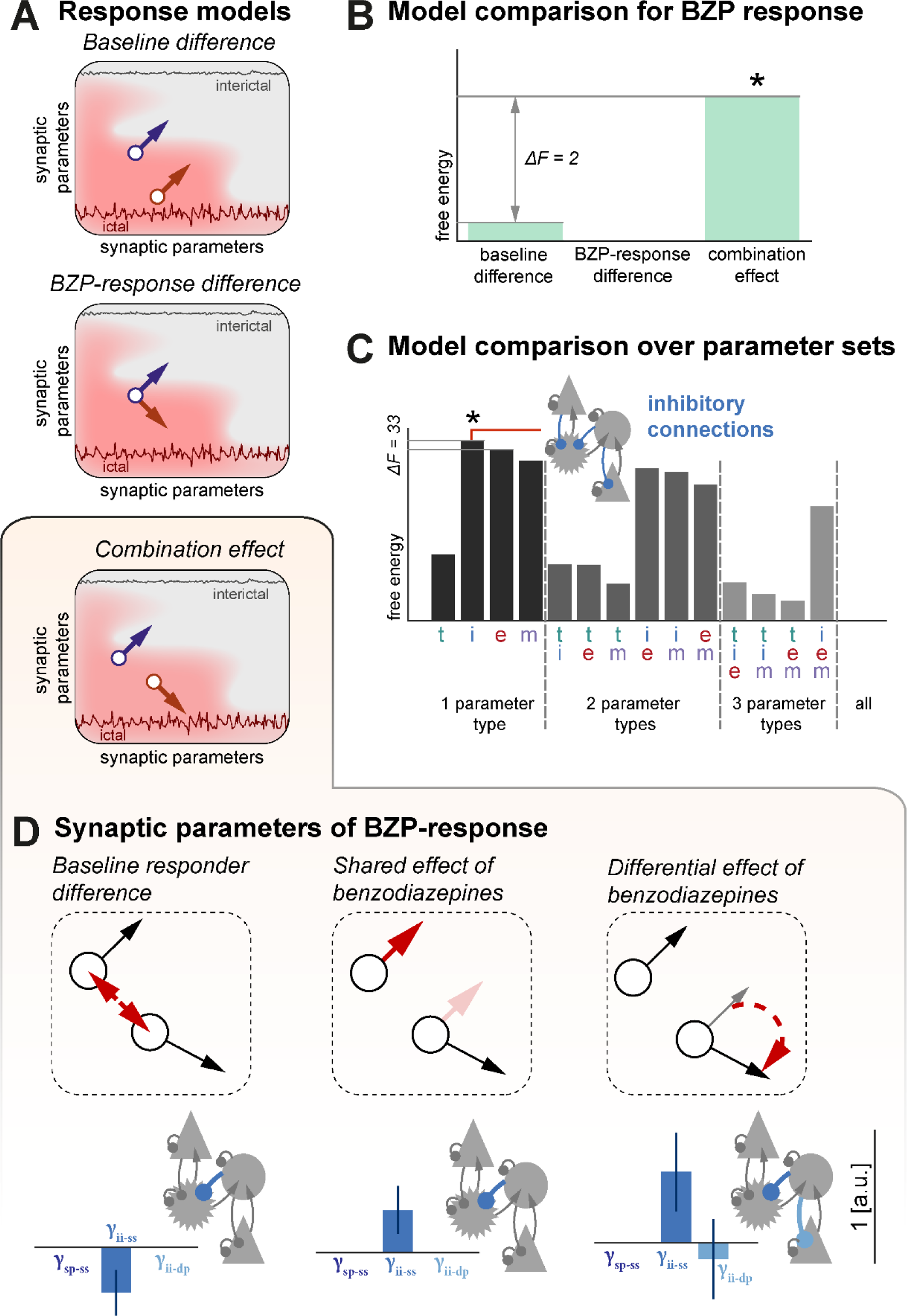
Benzodiazepine response is determined by both inherent and acquired differences in inhibitory synaptic coupling. (**A**) Models space for the difference between responders and non-responders, where their synaptic parameters; (top panel) differ during SE, but get similarly altered by BZPs; (middle panel) are similar during SE but are distinctly altered by BZPs; and (bottom panel) both differ in SE and are distinctly altered by BZPs. Across these models, the pre-medication state is indicated by a circle, the BZP effect is indicated by an arrow; orange signifies non-responders, purple signifies responders (**B**) Relative free energy calculated for each of the three hierarchical models revealing that the third, combined model best explains the variability in the EEG signals in our patient cohort. (**C**) Relative Free energy for a set of models in which this combined effect of BZP treatment across responders and non-responders is modelled only with subsets of microcircuit parameters, indicated as t – time constants, m – self-modulatory coupling, e – excitatory coupling, i – inhibitory coupling; multiple letters indicate combinations of these parameter sets. The model based on changes only in inhibitory coupling provided the best explanation for BZP treatment effects and the difference between responders and non-responders. (**D**) Synaptic parameters inferred in the winning model; inhibitory parameter values are shown for each of the constituent effects making up the winning from (**A**) where non-responders differ from responders during SE, and are differently altered by BZPs. This is described in three main constituent effects indicated in the red arrows: (i) a difference during established SE, (ii) a common response to BZP, and (iii) a selective response to SE in responders. These constituent effects are shown as red arrows, the full difference being inferred is shown as black circle and arrows. (Left panel, i) shows that responders have a lower level of GABA mediated inhibition onto spiny stellate cells during established SE (γ_ii-ss_); (middle panel, ii) shows that BZPs result in increased GABA-mediated inhibition on spiny stellate cells (γ_ii-ss_); and (right panel, iii) shows that in addition there is a further BZP-mediated increase in GABA mediated inhibition onto spiny stellate cells in the responders (γ_ii-ss_), coupled slight decrease in deep pyramidal cell inhibition (γ_ii-dp_). ‘*’, winning model (highest free energy); t, time constants,; m, self-modulatory coupling; e, excitatory coupling; i, inhibitory coupling; BZP, benzodiazepines; CMC, canonical microcircuit; DCM, dynamical causal modeling; GABA, γ-aminobutyric acid; PEB, parametric empirical bayes; PSD, power spectral density; vLFP = virtual local field potential.

To test these hypotheses, we inverted ‘second level’ models with subsets of the possible effects included in the full design matrix. When comparing the free energy for these models, the hypothesis 3 was the winning model. In a second step, we wanted to further test whether a subset of synaptic parameters is sufficient to capture the within and between subject effects. Models only allowing variation in inhibitory parameters showed the highest evidence (Fig. 3B). These findings suggest that the variability in the spectral modulation observed between BZP responders and non-responders in this cohort can be primarily attributed to the neural inhibitory drive (Fig. 3C-D, Supplementary Table 4).

### Probing of synaptic parameters reveals alternative mechanisms to terminate status epilepticus

The estimated hierarchical model identifies relevant contributions of microscale dynamics influencing the transition from SE to seizure termination following BZP administration. To validate our hierarchical model in delineating the conditions for state transition, we simulated power spectra mimicking the pathophysiological state prior to ASM administration in responders and non-responders and then navigated the parameter space along the shared and differential effects of BZP identified by the model. *In silico* state transition from the ictal to interictal state was observed only in responders, while no qualitative change was observed in non-responders (Fig. 4A). Based on a sensitivity analysis on the single synaptic parameter level (Supplementary Fig. 2), we identified alternative scenarios, where the shift in additional parameters suggests alternative, possible pathways for state transition also in non-responders (Fig. 4B), which are qualitatively distinct from those in responders.

**Figure 4:**
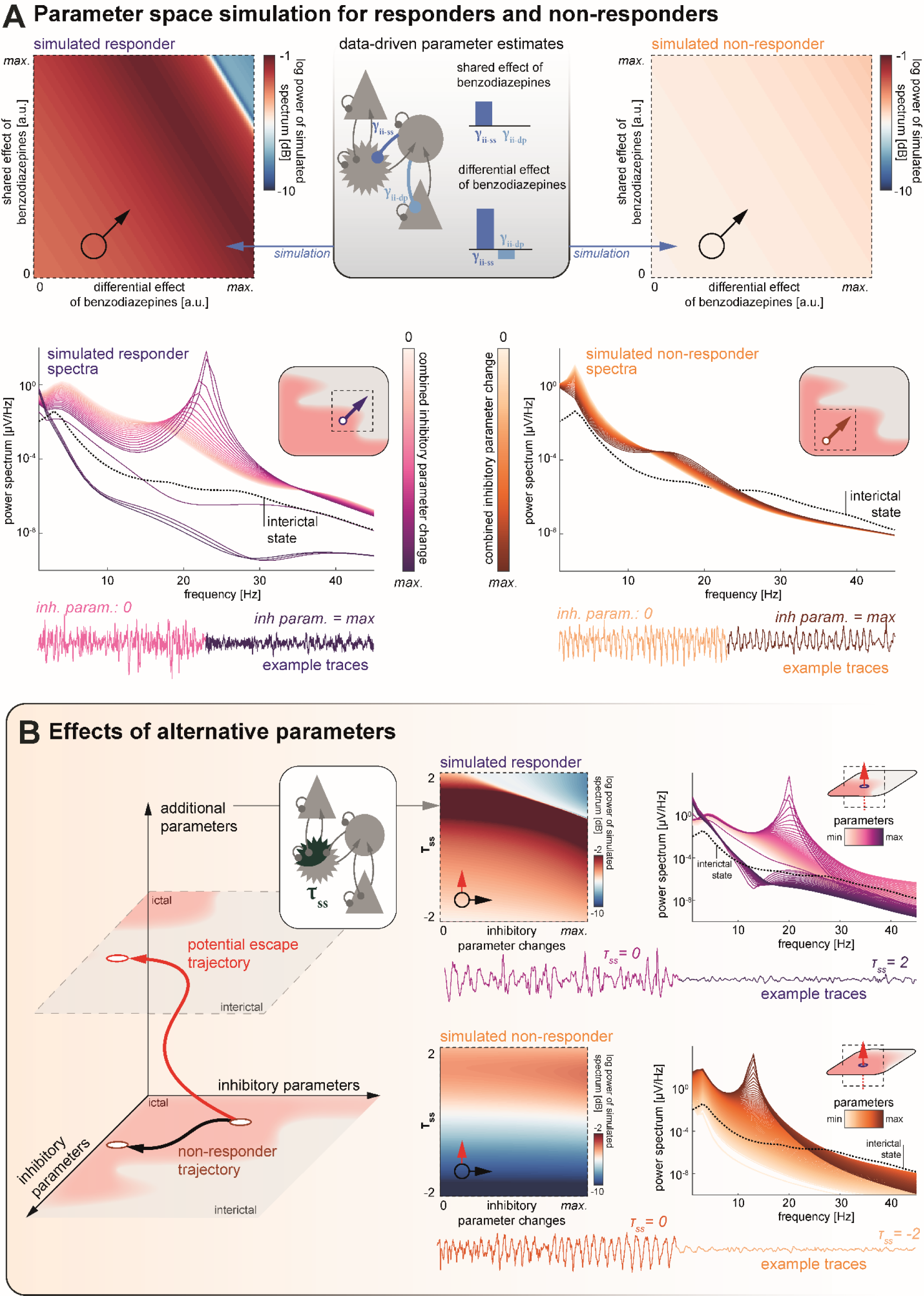
In silico probing of synaptic parameters reveals alternative mechanisms to terminate status epilepticus. (**A**) Power spectra (at 25 Hz) generated from a hierarchical parameter empirical Bayes (PEB) model (top panel, middle) demonstrating the relationship between the shared and differential response to BZPs and their combined effect on directing a transition from status epilepticus (SE, a continuous ictal state) towards termination (an interictal state). These simulations used parameters extracted from dynamic causal modelling (DCM) fitted to data from the responder (top panel, left) and non-responder (top panel, right) patient groups to reproduce changes in inhibitory coupling retrieved from dynamic causal modelling (DCM). A starting value was assigned to DCM parameters in responders and non-responders (BZP). Bottom panel, the power spectra along the diagonal trajectory (bottom left to top right, across inhibitory parameters and frequencies) from each of the corresponding power spectral plots in the top panel. The trace under the plot reflects the simulated EEG signal generated from the spectral properties observed. In the responder group, there is a discontinuity in the spectral power which corresponds with the transition from an ictal state (i.e., in SE) to an interictal state (i.e. SE termination) as shown in the insert. By contrast, in non-responders this transition is not present and reflects that these patients are further away from termination point (insert). (**B**) Schematic illustrating how this *in silico* approach to probing synaptic parameters can be used to reveal addition parameters that may contribute towards terminating SE such as the time constant of spiny stellate cells (τ_ss_, left panel). With the addition of τ_ss,_ it is possible to reproduce power spectra for the responders and non-responders by relating changes in τ_ss_ with changes in inhibitory parameters (middle and right panels). This demonstrates how manipulating τ_ss_ can push SE in both the responders and non-responders towards termination as shown in simulated EEG traces.

## Discussion

Our study provides evidence that pathological alterations of synaptic inhibition underlie BZP failure in paediatric SE. We demonstrated that spectral changes observed in clinical EEG following BZP administration are associated with the dynamic modulation of GABA-mediated inhibitory coupling in generative models of cortical dynamics. Furthermore, EEG spectral power showed two main effects in our cohort. First, responders to BZP treatment exhibited reduced low-frequency spectral power prior to treatment, which may indicate the electrographic signature of less prolonged SE. Second, responsiveness to BZPs was associated with a wideband decrease in neural synchronisation^23^, a well-documented effect following the potentiation of the GABAergic transmission.^24,25^

Our computational framework successfully captured the differences in excitatory-inhibitory coupling between responders and non-responders to BZP and detected a shift in inhibitory GABA-related drive following BZP administration. This is in line with experimental data suggesting that alterations in GABA-mediated inhibition underlie changes in BZP responsiveness.^7^ Our findings further demonstrate that this type of synaptic inhibition plays a crucial role in modulating the observed EEG dynamics in paediatric patients with SE, and further builds on the use of generative models to characterise synaptic mechanisms underlying EEG responses.^26^

The transition between dynamical states emerges from the slow fluctuation of non-linear evolving phenomena. Dynamical models have described the transition between seizures and physiological state in terms of critical transition and bifurcation analysis, inspired by and validated on experimental evidence.^27–29^ We successfully validated in-silico the role of the GABAergic drive in the transition to seizure termination. Notably, the enhancement of inhibitory coupling had a specific effect in a region of parameter space that aligned with the pre-medication EEG spectrum observed in responders, reproducing the spectral modulations typically observed after BZP administration.^30^ These findings underline the intricate relationship between microscale dynamics and macroscale brain activity, illustrating how subtle alterations in synaptic interactions can have profound effects on overall brain function in these complex dynamical systems. Importantly, generative models allow further interrogation of the dynamic landscape through simulations. Here, the computational framework showed alternative potential pathways in parameter space leading to SE termination, which were not observed in our dataset. In this context, DCM has the potential to prioritise potential alternative treatment strategies for empirical investigation and in future provide non-invasive monitoring for alternative therapeutic strategies.

Taken together, we provided compelling evidence that the effect of BZPs on macroscopic brain dynamics in paediatric patients with SE is primarily driven by dynamic shifts in inhibitory synaptic signalling. The observed inherent baseline variations could be leveraged through DCM to predict and optimise treatment responses in paediatric patients with SE.

## Data Availability

Raw EEG data is available upon reasonable request from the corresponding author. The code used to produce the results presented in this manuscript is available on an online repository.

https://github.com/tommytommy81/SE_DCM_code.

## Acknowledgements

We thank members of the University Children’s Hospital Zürich Neurophysiology Team for assistance in collection of patient data. We also thank Gerald Cooray (Great Ormond Street Hospital) for providing helpful comments on the manuscript. This study has received support from the Theodor & Ida Herzog-Egli Foundation Project (R.J.B., G.R., R.E.R.), the Anna Mueller Grocholski Foundation (T.F., R.J.B., G.R., R.E.R.), the Swiss National Science Foundation under grants agreement 208184 (G.R.), the Wellcome Trust (209164/Z/17/Z to R.E.R) and the European Union through the Human Brain Project / SGA3 (R.E.R.).

## Author contributions

Conceptualisation, T.F., R.J.B., G.R. and R.E.R.; Methodology, T.F. and R.E.R.; Investigation, T.F., R.J.B., A.S., G.S. and R.E.R.; Writing – Original Draft, T.F., R.J.B., G.R. and R.E.R.; Writing – Review and Editing, T.F., R.J.B., A.S., G.S., G.R. and R.E.R; Funding Acquisition, R.J.N., G.R. and R.E.R.

## Supplementary Methods

### EEG dimensionality reduction

We reduced the dimensionality of the data through an independent and identically distributed (IID) source localisation algorithm (SPM12) with the aim to capture each patient’s characteristic ictal rhythms in a single time series, analogous to previously published approaches.^1,2^ For this, representative 30s artefact-free ictal segments were visually identified from the EEG window prior to drug administration to generate an anatomical activation map that captures key cortical sources contributing significantly to ictal activity. Based on the cortical location with the highest amplitude activation we reconstructed the time course of the corresponding virtual Local Field Potential (vLFP) for the entire pre- and post-medication segments. This vLFP approach reconstructs signals at single points on the cortical mesh of a three-layer standardised head model using an empirical Bayes beamformer.^32^

### Dynamical Causal Modelling (DCM)

Briefly, DCM is a Bayesian model inversion framework in which parameters of dynamical models are fitted to time series data using a Variational Laplace approximation. Typically, modelling of oscillatory electrophysiological activity is performed by fitting synaptic coupling parameters of neural mass population models to cross spectral summaries of the presumed stationary time series.^31,33,34^

These model inversions yield posterior parameter estimates of the key synaptic coupling parameters, as well as a free energy estimate of the model evidence, which allows for subsequent comparison between models whilst balancing model accuracy and model complexity.^35^

### First level DCM

We then fitted spectral patterns in the vLFP before, and after drug administration using an adapted convolutional canonical microcircuit model within the DCM framework. Here, parameters are fitted for a model of a single cortical column consisting of four neuronal populations (superficial pyramidal cells, spiny stellate interneurons, inhibitory interneurons, deep pyramidal cells), which are coupled through recurrent excitatory and inhibitory connections.^16^ The dynamics of our adapted whole microcircuit are described by 14 key variable parameters that determine the time constants of their convolutive kernel (4), and the intrinsic excitatory (3), inhibitory (3) and self-modulating (4) coupling (Supplementary Tables 2 and 3). For each patient, two separate models were fitted – one parameterised to reproduce the pre-medication spectrum, one for the post-medication spectrum. These models were fitted to a broadband frequency spectrum (1-45 Hz) and adapted to suppress changes in non-neuronal scaling parameters within repeat inversions of models for the same patient.

### Second level DCM

The hierarchical modelling approach allows integrating a user-specified design matrix to link (1) ‘first level’ dynamic causal models that model separate neurophysiological datasets, e.g. data from individual patients, with (2) ‘second level’ hierarchical linear models that model the systematic variation of the first level model parameters, e.g. synaptic parameter differences between patient groups. These hierarchical models are inverted in a Bayesian framework that allows for propagating uncertainty over model parameters from the first to the second level, as well as optimisation of first level parameters based on iterative second level parameter estimates.^36,37^

We used the two first-level DCMs per patient that capture the pre- and post-medication window, and structured a second-level design matrix to estimate the across-DCM effects of whether the patient turned out to be a responder to BZPs (the ‘main effect of responsiveness’); the treatment with BZPs (the ‘main effect of benzodiazepines’) and an interaction term capturing conditional effects of BZPs specific to patients who ultimately respond (the ‘interaction term’). Treatment and interaction terms were included also for other ASM and considered as random effects. Details on the structure of the design matrix adopted for the hierarchical model inversion are provided in Supplementary Figure 2.

At the same time, we wanted to identify subsets of synaptic parameters which would provide the more parsimonious explanation for the observed differences both within-subject (pre- vs post-medication administration), and between-subjects (responders vs non-responders). For this we created a model space where only subsets of the synaptic model parameters were allowed to vary during the model inversion and divided parameters into functional classes (excitatory coupling, inhibitory coupling, modulatory recurrent self-connections, synaptic time constants), and comprehensively tested all combinations (i.e., 15 individual models). We then performed a fixed effects Bayesian model comparison across these second-level models, using the free energy approximation of the model evidence.^38^

### Simulations of effects of individual synaptic parameters

First, we identified initial conditions for the synaptic parameters by fitting a first level DCM to the average spectra of pre-medication responders and non-responders separately. Second, we systematically altered the value of synaptic parameters of choice, generating corresponding altered power spectra.

## Supplementary Information

**Supplementary Table 1:** Cohort of paediatric patients with status epilepticus. ‘Resp.’, Response to anti-seizure medication; ‘BAR’, barbiturate; ‘BZP’, Benzodiazepine; ‘LEV’, Levetiracetam; ‘PRO’, propofol; ‘VPA’, valproate

| Patient | Aetiology | Onset | Presentation | Type | Drug | Resp. |
| --- | --- | --- | --- | --- | --- | --- |
| 1 | genetic | generalized | subclinical | continuous | BZP | yes |
| 2 | genetic | generalized | convulsive | continuous | BZP | yes |
| 3 | genetic | focal | subclinical | continuous | BZP | yes |
| 4 | genetic | focal | subclinical | continuous | BZP | yes |
| 5 | metabolic | generalized | convulsive | continuous | BZP | yes |
| 6 | structural | focal | convulsive | continuous | BZP | yes |
| 7 | infectious | focal | subclinical | continuous | BZP | yes |
| 8 | autoimmune | focal | subclinical | intermittent | BZP | yes |
| 9 | genetic | generalized | subclinical | continuous | BZP | no |
| 10 | genetic | focal | subclinical | continuous | BZP | no |
| 11 | genetic | focal | subclinical | continuous | BZP | no |
| 12 | genetic | generalized | subclinical | continuous | BZP | no |
| 13 | genetic | generalized | subclinical | continuous | BZP | no |
| 14 | genetic | generalized | subclinical | continuous | BZP | no |
| 15 | metabolic | focal | subclinical | intermittent | BZP | no |
| 16 | genetic | generalized | subclinical | intermittent | BZP | no |
| 17 | autoimmune | generalized | subclinical | continuous | BZP | no |
| 18 | structural | generalized | convulsive | continuous | BAR | yes |
| 19 | autoimmune | generalized | convulsive | intermittent | BAR | yes |
| 20 | autoimmune | focal | subclinical | continuous | BAR | yes |
| 21 | infectious | focal | subclinical | intermittent | BAR | yes |
| 22 | structural | focal | convulsive | continuous | BAR | yes |
| 23 | structural | focal | convulsive | continuous | LEV | yes |
| 24 | structural | focal | convulsive | continuous | PRO | yes |
| 25 | structural | generalized | convulsive | intermittent | LEV | no |
| 26 | genetic | generalized | subclinical | continuous | VPA | no |

**Supplementary Table 2:**
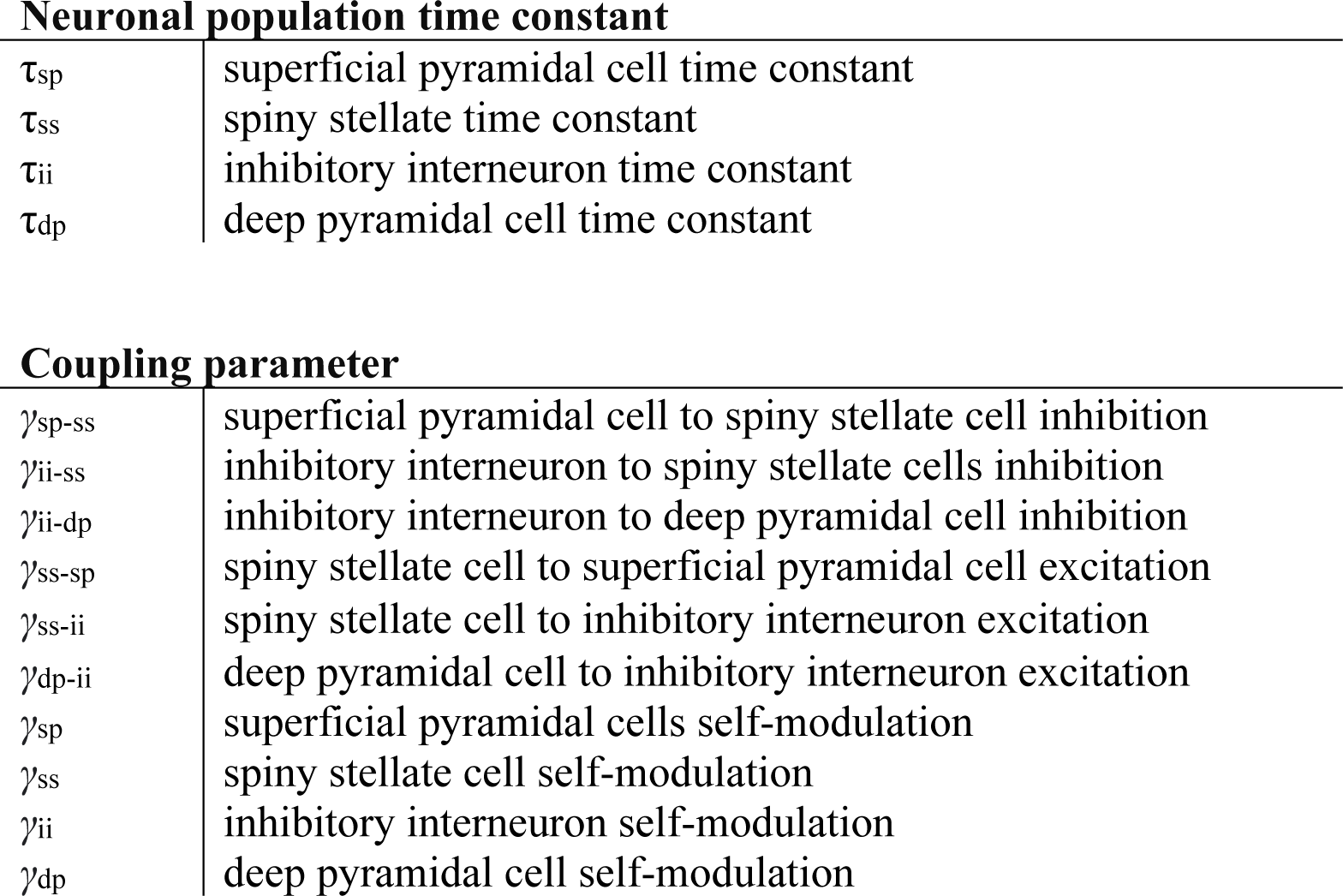
DCM parameters. List of synaptic parameters characterizing the CMC, which are fitted by the DCM. ‘CMC’, Canonical Micro-Circuit; ‘DCM’, Dynamical Causal Modelling.

**Supplementary Table 3:** CMC state equations. ‘CMC’, Canonical Micro-Circuit.’γ’, synaptic coupling (see Supplementary Table 2), ’τ’, synaptic time constant (see Supplementary Table 2), ‘S’, firing rate, ‘V’, membrane potential.

|  |  |
| --- | --- |
| $x_1$ | voltage (spiny stellate cells) |
| $x_2$ | conductance (spiny stellate cells) |
| $x_3$ | voltage (superficial pyramidal cells) |
| $x_4$ | conductance (superficial pyramidal cells) |
| $x_5$ | voltage (inhibitory interneuron) |
| $x_6$ | conductance (inhibitory interneuron) |
| $x_7$ | voltage (deep pyramidal cells) |
| $x_8$ | conductance (deep pyramidal cells) |
| $S_v$ | Firing rate dependent by membrane potential V |

**Supplementary Table 4:** Bayesian model comparison of second-level hierarchical PEB models. Free energy values the full factorial model space coupling the three hypothesized scenarios coupled with each of the 15 subsets of synaptic parameters. ‘t’, time constants; ‘m’, self-modulatory coupling; ‘e‘, excitatory coupling; ‘i’, inhibitory coupling; ‘PEB’, parametric empirical Bayes.

|  | Hypothesis/ Hierarchical PEB model |  |  |
| --- | --- | --- | --- |
|  | Baseline difference | BZP response difference | Combination effect |
| **t** | -19905.75 | -19901.58 | -19898.32 |
| **i** | -19476.52 | -19476.79 | **-19474.63** |
| **e** | -19508.47 | -19511.46 | -19508.08 |
| **m** | -19551.75 | -19559.56 | -19550.06 |
| **t i** | -19944.82 | -19940.18 | -19934.55 |
| **t e** | -19950.64 | -19944.78 | -19936.32 |
| **t m** | -20020.32 | -20016.07 | -20007.05 |
| **i e** | -19585.63 | -19591.31 | -19577.45 |
| **i m** | -19603.12 | -19612.06 | -19592.06 |
| **e m** | -19647.84 | -19658.49 | -19638.66 |
| **t i e** | -20025.23 | -20019.06 | -20002.64 |
| **t i m** | -20070.24 | -20068.47 | -20046.49 |
| **t e m** | -20095.44 | -20091.64 | -20070.30 |
| **i e m** | -19749.98 | -19765.42 | -19719.19 |
| **t i e m** | -20190.90 | -20194.63 | -20144.24 |

**Supplementary Figure 1:**
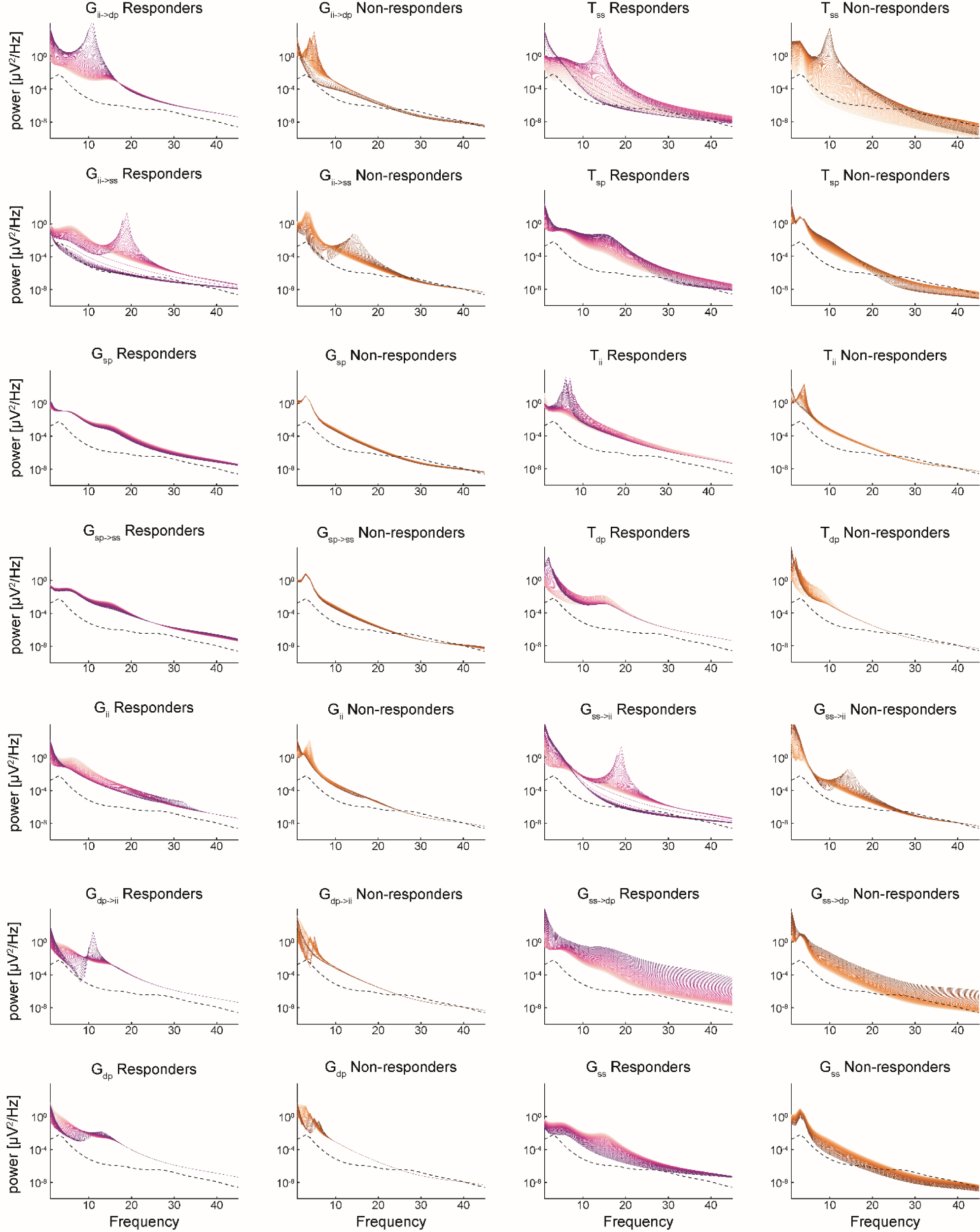
Sensitivity analysis. Results of sensitivity analysis across conductance (G) and time constant (T) parameters for each component of the cortical microcircuit. ‘ii’, inhibitory interneuron; ‘dp’, deep pyramidal; ‘sp’, superficial pyramidal; ‘ss’, spiny stellate.

**Supplementary Figure 2:**
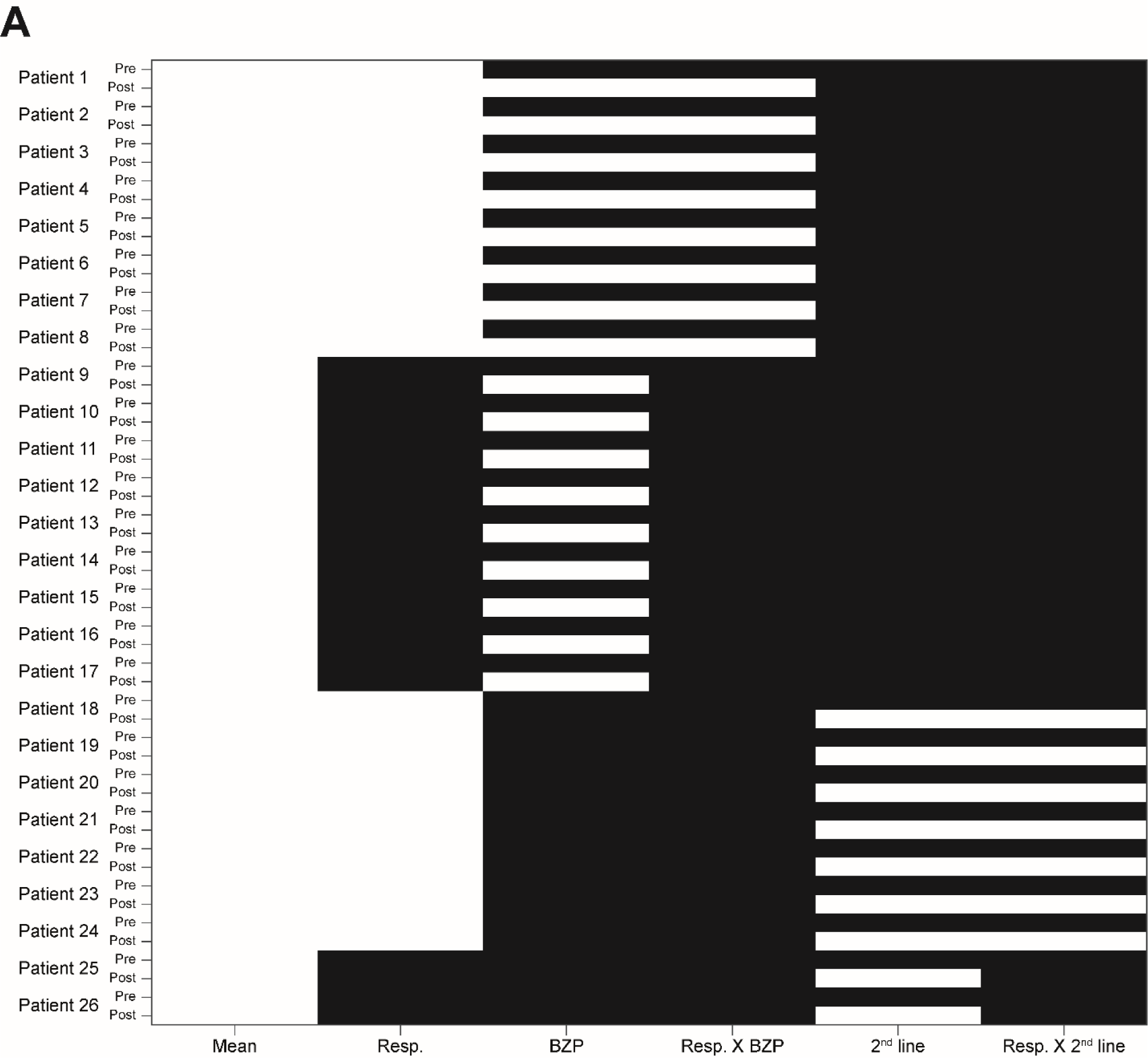
Design matrix used for hypothesis creation. Hypothesis 1 was modelled with a design matrix comprising the main effect of responsiveness and the main effect of BZPs; hypothesis 2 was modelled with a design matrix comprising the ‘main effect of BZPs’ and the ‘interaction term’; and hypothesis 3 was modelled with a full complement of the ‘main effect of responsiveness’, the ‘main effect of BZPs’ and the ‘interaction term’. ‘BZP’, benzodiazepine; ‘Resp’, response.

